# Cross-state variation in opioid use disorder among a privately insured nonelderly population in the United States

**DOI:** 10.1101/2020.06.09.20121269

**Authors:** Bibo Jiang, Li Wang, Douglas Leslie

## Abstract

**Background:** Although cross-state variation of the opioid epidemic in the United States are well documented in general, little are known about the epidemic in privately insured individuals.

**Objectives:** To describe cross-state variation in Opioid Use Disorder (OUD) among privately insured individuals in the US for the years 2005-2015 and investigate demographic differences of OUD patients between a group of hard-hit states and the rest states.

**Methods:** The MarketScan^®^ Commercial Claims and Encounters database was used to calculate prevalence of opioid use disorder for the 50 states in the US, respectively. We analyzed level and change of OUD prevalence in each state from 2005 to 2015 and identified the states which were affected most by the epidemic. One-sided exact fisher test was used to analyze demographic difference of the epidemic in the hard-hit states and the remaining states.

**Results:** Cross-state variations of the opioid epidemic among privately insured population were substantial, both in terms of severity and acceleration of the epidemic. Demographic patterns of the epidemic were similar across states. The 18-34 age group was the most affected group with the highest prevalence. The 55-64 group experienced the most rapid increase of OUD prevalence, especially in states that suffered most in the epidemic.

**Conclusions:** Results can assist policy makers to design better clinical and policy interventions on the opioid epidemic, especially on privately insured individuals. Drastic increase of OUD prevalence among the 55-64 group might indicate the need to improve prescription drug monitoring programs for chronic pain, especially in states more affected by the epidemic.

## 1. Introduction

In the past two decades, the opioid overdose epidemic has been one of the most pressing public health issues in the United States. Drug overdose deaths nearly tripled from 1999 to 2014, and over half of the deaths in 2014 involved an opioid.^2-5^ Recent statistics reveal a worsening of the opioid overdose epidemic. Opioid involved overdose death rate further increased by 15.6% from 2014 to 2015^3,6^ and 27.9% from 2015 to 2016^6^, resulting in 42,249 deaths (13.3 per 100,000 population) in 2016^6^. In addition to the rising rate of opioid involved mortality, the economic burden associated with opioid overdose, misuse and dependence is also increasing, reaching $78.5 billion in 2013.^7^

The magnitude of the crisis and the degree of adverse health and social outcomes have drawn particular attention from state and federal governments in recent years. In 2014, Massachusetts first declared the opioid epidemic a public health emergency, followed by Virginia in 2016 and Alaska, Arizona, Florida and Maryland in 2017.^8^ The federal government declared the opioid epidemic a national public health emergency on October 16, 2017. However, despite government efforts to curb the spread of the epidemic, opioid involved death rates have shown no signs of decreasing. The ongoing and increasingly severe opioid epidemic makes it crucial to closely track opioid overdose and understand how opioid use disorder risk varies across different subpopulations, such as different ethnic groups, age groups, gender and geographic location.

Although the opioid epidemic is nationwide, previous studies have documented geographic imbalances of the epidemic based on opioid-involved mortality rates across states, with the largest rates and increases concentrated in eastern states.^9-12^ A recent report released by the State Health Access Data Assistance Center confirmed that substantial differences in the opioid epidemic exist across states based on both non-heroin opioid death rates and heroin death rates. More specifically, in 2015, non-heroin opioid death rates ranged from 2.8 to 29.4 (per 100,000) and heroin death rates ranged from 0.7 to 13.3 (per 100,000) across states.^13^ Although geographic patterns in death rates associated with drug poisoning have largely been unexplored^10^, some empirical studies have suggested that the increase in death rates associated with drug poisoning has been greater for nonmetropolitan or rural areas of the U.S. as compared to metropolitan areas.^14,15^ Moreover, variation in the availability of opioid analgesics was found to be related to difference of drug poisoning mortality by state.^11,16^

Understanding variations in the opioid epidemic across states can assist state governments in designing localized interventions to more effectively address the epidemic. While previous studies have investigated demographic characteristics of the opioid epidemic in general, little is known about these patterns in privately insured populations. Since it has been suggested that a major cause of the epidemic is increased access to opioid pain medications in insured populations^17^, examining these patterns in privately insured population is important. Moreover, most previous studies about the opioid epidemic were on opioid overdose death rates using mortality data from death certificates, such as studies conducted by Centers for Disease and Control (CDC). However, due to the increasing heath care and economic burdens associated with OUD and its morbidities, studying prevalence of opioid use disorder is also very important to better understand the epidemic and help federal and state governments to allocate medical resources more effectively for OUD treatment and help reduce drug overdose death rates.

Medicaid plays a major role in financing treatment for opioid use disorder^18^; given its role as a safety net for individuals with OUD, this is not surprising. However, state variation in OUD prevalence in privately insured populations is not well understood. The present study sought to fill the existing gap by investigating state-level OUD prevalence using data from the MarketScan® Commercial Claims and Encounters database for the period from 2005 to 2015.

## 2. Methods

### 2.1 Data Source

Data used in this study are from the MarketScan^®^ Commercial Claims and Encounters database (MarketScan), which includes claims information from various payers, and describes the health care service use and expenditures for covered employees and family members. The database includes subsections, including inpatient claims, outpatient claims, outpatient prescription drug claims, and enrollment information. Claims data in each of the subsections contain a unique encrypted patient identifier and information on patient age, sex, geographic location (e.g., state), etc. The unique encrypted patient identifier, which is not available in admission-based data systems such as TEDS, makes it feasible to count the number of OUD patients for each state in each year.

### 2.2 Study Sample

The MarketScan data we used in this study covers the period from January 2005 through December 2015 for all 50 states and contains health care service information for tens of millions of privately insured individuals each year (varying from year to year). The sample includes approximate 20 to 50 million individuals each year, and does not contain any individuals aged 65 or older. Therefore, the analysis is on a privately insured non-elderly (aged 64 or younger) cohort. The demographic characteristics stay relatively stable over this period. Institutional review board approval was obtained prior to implementation of the study.

### 2.3 Study Measures

Opioid use disorder is a diagnosis introduced in the fifth edition of the Diagnostic and Statistical Manual of Mental Disorders (DSM-5). It combines two disorders from the previous edition of the Diagnostic and Statistical Manual, the DSM-IV-TR, known as Opioid Dependence and Opioid Abuse, and incorporates a wide range of illicit and prescription drugs from the opioid class. Following previous studies^19^, we identified patients with OUD as those with an inpatient or outpatient claim with a primary ICD-9 diagnosis code of 304.0x, 304.7x, 305.5x, 965.0x, E850.0 – E850.2, or E935.0 – E935.2 and excluded those with a diagnosis of self-inflicted poisoning (E950.0 – E950.5) and assault by poisoning (E962.0x). Beginning in October 2015, ICD-10 diagnosis codes instead of ICD-9 diagnosis codes were used. Accordingly, for claims occurring in October 2015 and forward, we used ICD-10 code F11.xx and T40.0 –T40.6 to identify OUD patients, and excluded those with a diagnosis of self-poisoning (T40.0X2, T40.1X2, T40.2X2, T40.3X2, T40.4X2, T40.5X2 and T40.6X2) and assault-by-poisoning (T40.0X3, T40.1X3, T40.2X3, T40.3X3, T40.4X3, T40.5X3 and T40.6X3).

State level OUD prevalence rates were calculated using the total number of OUD patients divided by the total number of enrolled individuals for that year in each state. Because there are no enrolled individuals aged 65 or older, the OUD prevalence rate is essentially for a non-elderly cohort (aged 64 or younger) only. OUD prevalence in 2015, the ending year of the sample period was investigated to study same-year cross-state variations. Moreover, absolute changes and relative changes in prevalence rates^2^ between the two years were also calculated to study different trends of evolution of the epidemic across states.

Due to diagnosis codes switching from ICD-9 to ICD-10 in October 2015, the prevalence rates calculated for 2015 are not directly comparable to the prevalence rates calculated for 2005. To adjust for the effect from diagnosis codes switching, we assume that the number of OUD patients identified in the first three quarters is a fixed fraction of the total number of OUD patients identified throughout the year. The data largely supports the assumption since this fraction was 84.4% in 2013 and 84.7% in 2014. However, this fraction became 81.2% in 2015, which indicates that the number of the fraction of OUD patients identified in the fourth quarter of 2015 was higher than the previous years. This was likely due to the switching of diagnosis codes. Based on this assumption, we have:

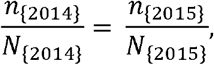

where *n*_{2014}_ and *n*_{2015}_ denote the number of OUD patients identified in the first three quarters of 2014 and 2015 respectively and *N*_{2014}_ and *N*_{2015}_ denote the total number of OUD patients identified in the corresponding years. Therefore, we calculate the adjusted number of OUD patients in 2015 using the following formula:

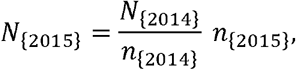

and calculate adjusted OUD prevalence rate for 2015 using the adjusted number of OUD patients and the total number of enrollments in 2015. In the following analysis, all OUD prevalence rates in 2015 were adjusted using this method.

### 2.4 Data Analysis

We use US maps in Figure 2 and Figure 3 to show variations of the opioid epidemic across states. Figure 2 shows variation of OUD prevalence across states in 2015. The OUD prevalence rates for the 50 states were summarized into 5 ranges such that each range contains 10 states. To show different trends of evolution for the epidemic across states, we present absolute and relative changes of OUD prevalence rates from 2005 to 2015 in Figure 3.

To gain more insight on the opioid epidemic in the states that suffer most from the epidemic, we classified the 50 states into two groups based on OUD prevalence rates in 2015. The 10 states with the highest OUD prevalence in 2015 were classified as the state group experiencing a relatively severe opioid epidemic and the remaining states were used as a comparison group. To investigate contributing groups to the relatively high OUD prevalence rates in these 10 states, we calculated the OUD prevalence in subpopulations identified by age group and gender, and compared those with their corresponding counterparts in the remaining states where the opioid epidemic was less severe. The relative change in the prevalence between 2005 and 2015 was also calculated for each group in each state to identify different rates of increase. Five age groups were considered as follows: 0-17,18-35, 35-44, 45-54, and 55-64.

We applied a one-sided fisher exact test on the OUD prevalence to compare demographic differences between the 10 states with more severe opioid epidemic and the remaining states. The analysis was conducted using SAS 9.4.

## 3. Results

### 3.1 The Opioid Epidemic Across States

As shown in Figure 1, national level of OUD prevalence had monotonically increased from 2005 to 2015. It reached 222.78 per 100,000 population in 2015, more than 5 times of the level in 2005 (39.48 per 100,000). Virtually all states experienced dramatic increase on OUD prevalence during this period. In 2005, the median OUD prevalence in the 50 states was 40.4 (per 100,000) and all states except for Michigan (112 per 100,000) had OUD prevalence rates below 100 per 100,000. However, in 2015, only four states including South Dakota, Nebraska, Iowa and Hawaii had OUD prevalence rates below 100 per 100,000 population. The median OUD prevalence in the 50 states reached 239.3 (per 100,000), which was almost 6 times the median level in 2005.

**Figure 1:**
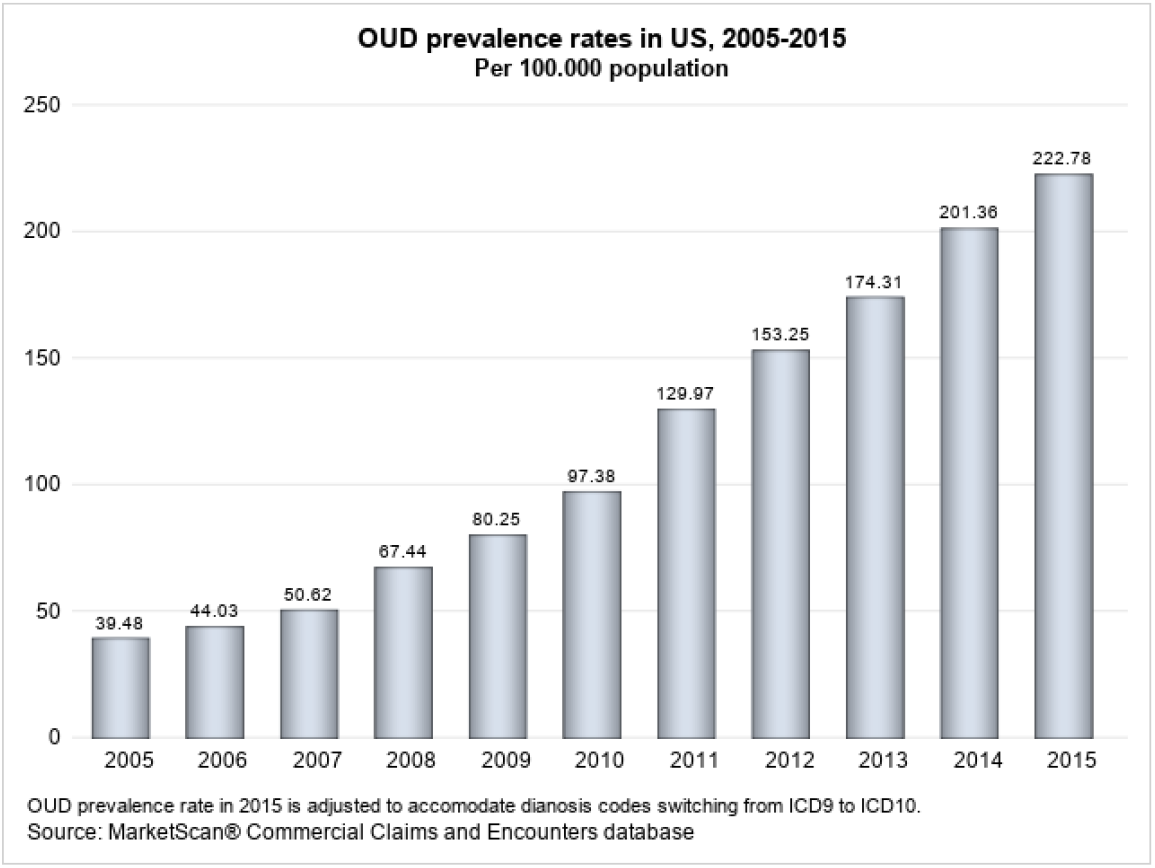
National OUD prevalence rates from 2005 to 2015

Although all the 50 states experienced fast increase on OUD prevalence, we can tell from Figure 2 that the level of prevalence rates vary substantially across states. The prevalence rates of the 50 states are categorized into 5 ranges, with the highest range being 337.8 to 542.0 per 100,000, and the lowest range being 57.3 to 133.7 per 100,000. The Eastern United Sates suffered most from the opioid epidemic in 2015 and the Central United States had relatively low OUD prevalence rates. West Virginia, Rhode Island, Tennessee, Maine, Delaware, Washinton State, New Hampshire, Vermont, Connecticut and Louisiana were the 10 states with the highest OUD prevalence rates in 2015. Prevalence rates of these 10 states varied from 337.8 to 542.0 per 100,000 population, much higher than those states in the Central United States, such as South Dakota (57.3), Nebraska (84.8), Iowa (85.7), Hawaii (98.6), Minnesota (105.9) and Kansas (111.9). The epidemic in the Western United States in general was not as severe as that in the Eastern United States. However Washington State (386.4), Utah (276.7) and Alaska (271.3) were the three states in this region that had OUD prevalence rates among the top 20 states.

**Figure 2:**
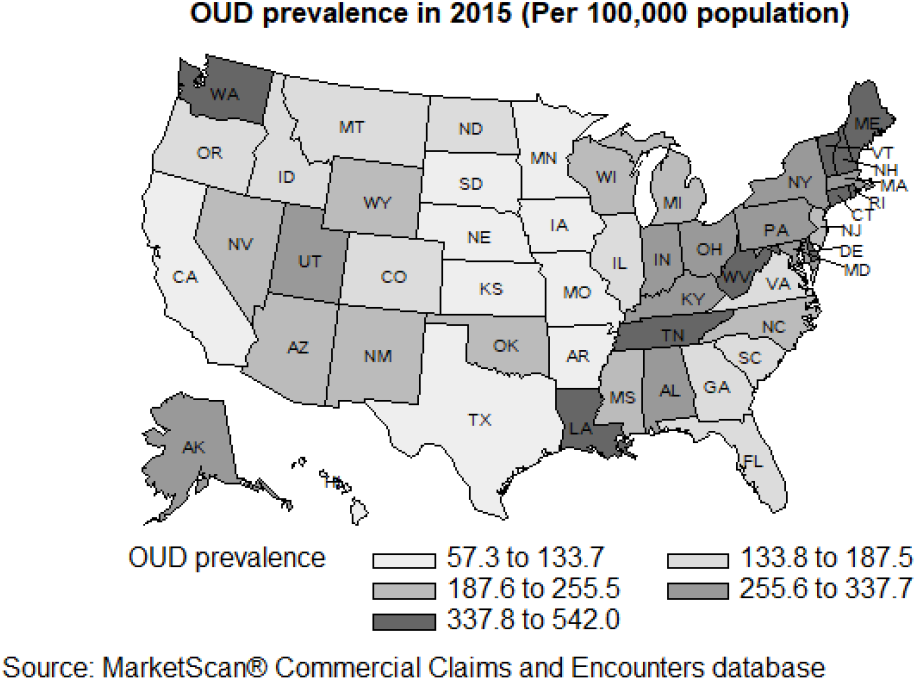
OUD prevalence rates by state (2015)

To study the trends of evolution of the epidemic in the 50 states, we present absolute change^3^ and relative change^4^ of the state-level OUD prevalence rates during the 11-year timeframe in Figure 3. From the map for “Absolute changes”, we can see that in general those suffered more from the epidemic in 2015 were also the ones that had big absolute changes on prevalence rates from 2005 to 2015. Most of the Eastern United states had OUD prevalence increase by 227.4 to 473.2 per 100,000 population, while the increase on the prevalence rates in the Central United States mostly varied within the range of 43.3 to 150.7 per 100,000. On the other hand, the map for “Relative changes” showed cross-state variation on percentage change of the prevalence rate during this period. The Eastern United States again showed higher increasing rates than most states in other regions. To conclude, most states that suffered most in 2015 showed rapid increase on the prevalence rate both in terms of absolute and relative changes. Another finding worth of noting is that Wyoming and New Mexico had the most rapid increase in terms of relative change, with Wyoming increasing by 1002% (from 17.8 to 188.3 per 100,000) and New Mexico increasing by 984.5% (from 21.21 to 230.1 per 100,000). Although the high relative changes of the prevalence rates in these two states were partially attributable to the low prevalence rates in the two states in 2005, the rates of increase were still astonishing. Some other states in the same region, such as Oklahoma, Colorado and Missouri also had rapid increase on OUD prevalence, with increasing rate being 546.2%, 477.9% and 453.9%, higher than the median increasing rate 378.9%.

**Figure 3:**
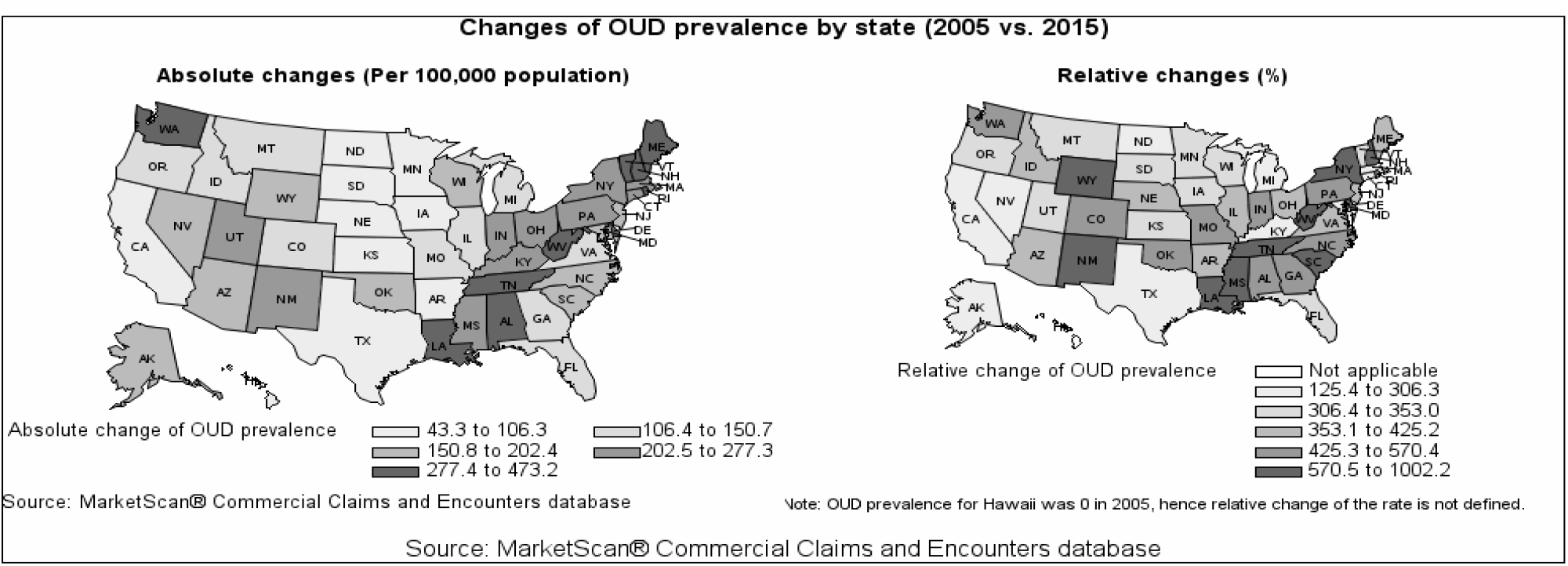
Absolute and relative changes of OUD prevalence rates by state (2005 vs. 2015)

### 3.2 Opioid Epidemic Development in Hard-hit States

As mentioned in the previous section, West Virginia, Rhode Island, Tennessee, Maine, Delaware, Washinton State, New Hampshire, Vermont, Connecticut and Louisiana were the 10 states with the highest OUD prevalence rates in 2015, hereafter collectively referred as “T10” states. These states also experienced faster increase on OUD prevalence rates than most other states did. Because of the more severe epidemic in these states, we further analyze trends of evolution of the epidemic in T10 states by analyzing level and change of OUD prevalence in subpopulations defined by gender and age groups.

Table 1 lists the ranks of T10 states based on state-level OUD prevalence rates in each year. It shows that the majority of T10 states had ranked high in OUD prevalence since 2005. With the exception of Louisiana, T10 states had remained within the top 20 states for OUD prevalence in most years during the 11-year timeframe. Louisiana, which was ranked the 10^th^ highest for OUD prevalence in 2015, had ranked within the 20-30 range from 2005 to 2013, but experienced a rapid increase in 2014 and 2015, resulting in its 10^th^ position in 2015.

**Table 1:** Historical Ranks of OUD Prevalence for T10 States

| State Name | 2005 | 2006 | 2007 | 2008 | 2009 | 2010 | 2011 | 2012 | 2013 | 2014 | 2015 |
| --- | --- | --- | --- | --- | --- | --- | --- | --- | --- | --- | --- |
| West Virginia | 9 | 11 | 1 | 1 | 1 | 1 | 1 | 1 | 1 | 2 | 1 |
| Rhode Island | 11 | 9 | 6 | 2 | 2 | 2 | 2 | 2 | 2 | 1 | 2 |
| Tennessee | 13 | 17 | 14 | 16 | 15 | 14 | 15 | 15 | 17 | 8 | 3 |
| Delaware | 19 | 25 | 23 | 8 | 4 | 8 | 3 | 3 | 3 | 5 | 4 |
| Maine | 6 | 8 | 10 | 10 | 11 | 5 | 5 | 5 | 5 | 7 | 5 |
| Washington | 12 | 14 | 7 | 4 | 5 | 7 | 6 | 7 | 8 | 6 | 6 |
| Vermont | 1 | 2 | 4 | 2 | 3 | 4 | 10 | 6 | 7 | 3 | 7 |
| New Hampshire | 28 | 13 | 15 | 19 | 18 | 12 | 14 | 12 | 6 | 9 | 8 |
| Connecticut | 3 | 3 | 3 | 5 | 7 | 4 | 10 | 6 | 7 | 3 | 9 |
| Louisiana | 21 | 26 | 19 | 22 | 27 | 23 | 21 | 23 | 22 | 15 | 10 |

Apart from being historically high, OUD prevalence in T10 states in general increased more rapidly when compared to the remaining states^5^, hereafter collectively referred as “RM” states. The increasing rates of OUD prevalence in eight of the T10 states were above the median of 387%. Among them, seven exceeded 500%. Maine and Connecticut were the only two states in T10 states that had increasing rate being 307.9% and 302.6% respectively, lower than the median increasing rate. However, the relative lower increasing rates in the two states are partially attributable to their high prevalence rates in 2015. The prevalence rate for Vermont and Connecticut was the 2^nd^ and 3^rd^ highest in 2015. The absolute change of OUD prevalence in these two states were 288.3 and 265.3 per 100,000 population, much higher than the median level 174.1 per 100,000 population. Therefore, it is reasonable to conclude that the 10 states that suffered most from the epidemic in 2015 also experienced rapid increase on OUD prevalence during the 11-year timeframe.

### 3.3 Demographic Difference of the Opioid Epidemic Between Two State Groups

To further examine the fast spread of the opioid crisis in T10 states, we investigated the level and change of OUD prevalence in various subpopulations in T10 states. In particular, we analyzed OUD prevalence in subpopulations classified by gender and age groups. Table 2 lists the OUD prevalence in gender and age groups for T10 states in 2005 and 2015. As a comparison, the corresponding prevalence rates for RM states are also shown. To test whether the OUD prevalence rate for each subpopulation in T10 states is significantly higher compared to the corresponding subpopulations in RM states, we applied the one-sided exact fisher test and presented the p-values in Table 2 as well.

**Table 2:** OUD Prevalence (per 100,000) of Subpopulations and Relative Change of OUD Prevalence from 2005 to 2015

|  | 2005 |  |  | 2015 |  |  | % change |  |
| --- | --- | --- | --- | --- | --- | --- | --- | --- |
|  | T10 | RM | p-value | T10 | RM | p-value | T10 | RM |
| Age group |  |  |  |  |  |  |  |  |
| 0-17 | 13.9 | 10.3 | 0.010 | 17.3 | 13.1 | 0.002 | 24.5 | 27.2 |
| 18-34 | 112.9 | 64.7 | <0.001 | 636.5 | 365.4 | <0.001 | 463.7 | 464.8 |
| 35-44 | 82.3 | 60.2 | <0.001 | 527.0 | 263.4 | <0.001 | 540.3 | 337.5 |
| 45-54 | 72.7 | 66.8 | 0.090 | 409.0 | 207.5 | <0.001 | 462.6 | 210.6 |
| 55-64 | 36.6 | 33.3 | 0.183 | 327.2 | 179.7 | <0.001 | 794.0 | 439.6 |
| Sex |  |  |  |  |  |  |  |  |
| Male | 73.3 | 51.3 | <0.001 | 454.0 | 252.6 | <0.001 | 519.4 | 392.4 |
| Female | 53.3 | 39.0 | <0.001 | 308.9 | 158.7 | <0.001 | 479.5 | 306.9 |

According to Table 2, the patterns of OUD prevalence across subpopulations were similar in the two state groups, T10 states and RM states. From 2005 to 2015, both state groups experienced a dramatic increase in OUD prevalence for all age groups with the exception of the 0-17 age group. Younger groups (except for the 0-17 age group) generally had higher OUD prevalence rates than older groups, with the peak prevalence appearing in the 18-34 age group. It is worth noting that in RM states, the OUD prevalence rate in the 45-54 age group was the highest across all age groups in 2005, but was lower than that in the 18-34 and 35-44 age groups in 2015, indicating that the epidemic in the two younger groups deteriorated much faster than the age group 45-54 in RM states.

Although the patterns of OUD prevalence across different subpopulations were similar, the difference between the two state groups was apparent. OUD prevalence level for each subpopulation was generally much higher in T10 states than in RM states for both years, with the exception that the difference of the prevalence rates between the two state groups in both 45-54 and 55-64 age groups was either marginally (p-value = 0.090) or not (p-value= 0.183) significant in 2005. However, the difference of the prevalence rates in these two age groups became statistically significant between the two state groups in 2015.

Although Table 2 shows that the 18-34 age group had the highest OUD prevalence rate across all age groups in T10 states for both years, the 55-64 age group had the biggest increasing rate on OUD prevalence from 2005 to 2015, with a percentage change of 794%. Although the same age group in RM states also experienced a dramatic increase (439.5%) on OUD prevalence, the increasing speed was still much lower than that in T10 states. In fact, the relative changes of OUD prevalence for the 45-54 and 55-65 age groups in T10 states were about twice of those for the same age groups in RM states. These differences were much larger than observed in the other age groups.

In addition, Table 2 also shows that the change of OUD prevalence was drastic for both males and females in both state groups. Prevalence rates in both years and state groups were much higher in males compared to females, suggesting that males are likely more vulnerable to this disease than females.

## 4. Discussion

Regardless of the rapid increase in OUD prevalence nationwide, there is a significant difference in the OUD epidemic across states. The analysis results show that the Eastern United States suffered most from the epidemic in 2015. This finding is mostly consistent with CDC findings based on rates of drug overdose death by state in 2015^2^°.ln particular, West Virginia (41.5 per 100,000 population) was the one with the highest overdose death rate, following by New Hampshire (34.3). The overdose death rates for Rhode Island, Tennessee, Connecticut, Delaware and Maine varied from 21.2 to 28.2 per 100,000, much higher than the median overdose death rate 16.1 (per 100,000) of the 50 states in 2015. It is worth noting that the overdose death rate for Utah and New Mexico was 23.4 and 25.3 (per100,000) respectively, ranked among the top 10 according to the CDC report. However, the OUD prevalence rates for these two states ranked 17 and 24 among the 50 states based on our analysis results. On the other hand, Washington State, which had the 6^th^ highest OUD prevalence rate in 2015 based on our results, had overdose death rate being 14.7 (per100,000), lower than the median death rate among the 50 states. The difference between our results and the CDC report death rates might due to different research populations since our study sample is from a privately insured population while the CDC report was based on death certificates which were drawn from the whole population. Another possible reason for the difference might indicate different OUD treatment effectiveness across states. States with same OUD prevalence rate and more effective treatment programs are likely to have lower overdose death

Moreover, our results showed a staggering increase in the OUD prevalence rate in eastern states, most of which were historically harder-hit states as well. The rapid deterioration of the epidemic in these states indicate that the epidemic in these hard-hit states showed no signs to slow down. More effective interventions are needed in those states to curb the epidemic. In addition, our results also show a skyrocketing rate of increase in some Midwest states. This suggests that the OUD epidemic could be spreading to regions previously less impacted. More active and effective interventions should be taken in those states to prevent the rapid spread of the epidemic.

Previous studies found that the opioid epidemic did not conform to the stereotypical boundaries of age, gender, and geography. The young males had been the most affected group.^16,21^ Our study based on data from a privately insured population confirmed that almost every population subgroup was affected by the OUD epidemic, and the 18-34 age group appeared to be the most vulnerable group having the highest OUD prevalence across all the age groups. In addition, we found that after the mid-30s, increasing age was associated with a decrease in OUD prevalence. This is consistent with the previous finding that OUD prevalence peaks among those between 18 and 29 years of age and then decreases as age increases.^22^ This might partly be due to early mortality or remission of symptoms after reaching age 40.^23^ It is also worth noting that although our findings show that the prevalence of the disorder was highest in the 18-34 age group, OUD prevalence for the older group aged 55-64 increased most rapidly, especially in T10 states. More specifically, from 2005 to 2015, OUD prevalence increased 794.0% in T10 states and 439.6% in RM states for the 55-64 age group. This drastic increase of OUD prevalence in the 55-64 age group was likely caused by the dramatic increase of prescription opioids during this period, since this age group in general is more likely to use opioid involved pain medications than other groups. The fact that the increasing rate was much higher in T10 states might indicate more aggressive opioid prescribing activities in those states. A multifaceted approach including educating health professionals and the public about appropriate use of opioids, implementing prescription drug monitoring programs, enhancing the accessibility of nonopioid pain medications, etc. should be taken to battle against the disorder.

### 4.1 Limitation

The findings in this research are subject to the following limitations. Firstly, the data we used in the study cover the period from 2005 to 2015, which do not catch recent trends of evolution of the OUD epidemic. Analysis using more recent data would be meaningful. Secondly, the identification of OUD patients in this study was based on OUD related diagnosis codes from outpatient and inpatient claims. Because drug claims do not contain diagnosis codes, we were not able to identify those patients who only had claims for medications used to treat OUD (e.g., buprenorphine) for the entire year. From this point of view, the OUD prevalence calculated in this study might be underestimated. However, we believe that the bias may be minimal since clinical guidelines state that patients on OUD pharmacotherapy should be monitored regularly and receive psychosocial treatment^24^, which would result in inpatient or outpatient claims. Moreover, like all studies based on claims data, our study was not able to catch those OUD patients who never seek treatment. This is another possible factor causing underestimation of OUD prevalence in the commercially insured population. In addition, due to diagnosis codes switching from ICD-9 to ICD-10 in October 2015 and the fact that no one-on-one mapping exists between the two sets of codes, the classification of OUD patients might not be fully consistent. Although we have made reasonable adjustment on OUD prevalence in 2015, the estimation of OUD prevalence in 2015 and the change of prevalence from 2005 to 2015 can still be affected by the change of diagnosis codes.

## 5. Conclusions

By investigating state level OUD prevalence from 2005 to 2015 using claims data from the MarketScan, we found that although the opioid epidemic was nationwide and most states had experienced drastic increases in OUD prevalence, cross-state variations were also substantial, both in terms of severity and acceleration of the epidemic. The sharp increase of OUD prevalence and variations across states indicate that more effective and localized interventions are needed to curb the epidemic. The exceptionally drastic increase of OUD prevalence among the 55-64 age group might indicate the need to improve prescription drug monitoring programs, especially in states more affected by the epidemic. Our study provides a valuable reference for policy makers to design targeted clinical and policy interventions to address the opioid epidemic in the United States.

## Data Availability

The study used only secondary data from private insurance medical claims database (Marketscan). Although we are not allowed to share the data. The data are publicly available with purchase.

## Conflicts of interest

The authors report no conflicts of interest.

## Footnotes

2 Absolute change of prevalence refers to the simple difference in the prevalence rate over two periods in time; relative change of prevalence refers to percentage change of the prevalence rate over two periods in time.

3 Absolute change is defined as *P*_{2015}_ − *P*_{2005}_, where *P*_{2015}_ and *P*_{2005}_ represent OUD prevalence rate in 2015 and 2005, respectively.

4 Relative change is defined as 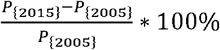.

5 Hawaii was excluded in this part of study since Hawaii had no OUD patients in 2005 and relative change of OUD prevalence from 2005 to 2015 was not defined mathematically.

## Notes

### Competing Interest Statement

The authors have declared no competing interest.

### Funding Statement

No external funding was received.

### Author Declarations

The study used only secondary data from private insurance medical claims database (Marketscan). The database has been stripped of identifiers. The research material obtained from human subjects is in the form of data acquired from IBM Truven Health Analytics, specially designed for research use. The subjects from the database that made available to us can be neither identifiable nor linkable by us.

## References

1. Bachhuber MA, Mehta PK, Faherty LJ, Saloner B. Medicaid Coverage of Methadone Maintenance and the Use of Opioid Agonist Therapy Among Pregnant Women in Specialty Treatment. Med Care. 2017;55(12):985–990.

2. CDC. Drug Poisoning Mortality: United States, 1999-2015. 2017.

3. Rudd RA, Seth P, David F, Scholl L. Increases in Drug and Opioid-Involved Overdose Deaths - United States, 2010-2015. MMWR Morb Mortal Wkly Rep. 2016;65(5051):1445–1452.

4. Rudd RA, Aleshire N, Zibbell JE, Gladden RM. Increases in Drug and Opioid Overdose Deaths— United States, 2000-2014. MMWR Morb Mortal Wkly Rep. 2016;64(50-51):1378–1382.

5. Prevention USCfDCa. Increases in Drug and Opioid-Involved Overdose Deaths — United States, 2010-2015.: Morbidity and Mortality Weekly Report. 2016.

6. CDC. Overdose Deaths Involving Opioids, Cocaine, and Psychostimulants — United States, 2015-2016. Morbidity and Mortality Weekly Report (MMWR) 2018;67(12):10.

7. Florence CS, Zhou C, Luo F, Xu L. The Economic Burden of Prescription Opioid Overdose, Abuse, and Dependence in the United States, 2013. Med Care. 2016;54(10):901–906.

8. Rutkow L, Vernick JS. Emergency Legal Authority and the Opioid Crisis. N Engl J Med. 2017;377(26):2512–2514.

9. Warner M, Chen LH, Makuc DM, Anderson RN, Minino AM. Drug poisoning deaths in the United States, 1980-2008. NCHS Data Brief. 2011(81):1–8.

10. Rossen LM, Khan D, Warner M. Trends and geographic patterns in drug-poisoning death rates in the U.S., 1999-2009. Am J Prev Med. 2013;45(6):e19–25.

11. Paulozzi LJ, Ryan GW. Opioid analgesics and rates of fatal drug poisoning in the United States. Am J Prev Med. 2006;31(6):506–511.

12. Seth P, Scholl L, Rudd RA, Bacon S. Overdose Deaths Involving Opioids, Cocaine, and Psychostimulants - United States, 2015-2016. MMWR Morb Mortal Wkly Rep. 2018;67(12):349–358.

13. Planalp CaL, Megan. The Opioid Epidemic: State Trends in Opioid-Related Overdose Deaths from 2000 to 2015. 2017.

14. McDonald DC, Carlson K, Izrael D. Geographic variation in opioid prescribing in the U.S. J Pain. 2012;13 (10):988–996.

15. Centers for Disease C, Prevention. CDC grand rounds: prescription drug overdoses - a U.S. epidemic. MMWR Morb Mortal Wkly Rep. 2012;61(1):10–13.

16. CDC. Vital Signs: Overdoses of prescription opioid pain relievers - United States, 1999-2008. MMWR Morb Mortal Wkly Rep. 2011;60(43):6.

17. Kounang N. 41 state attorneys general subpoena opioid manufacturers. CNN.2017.

18. Becker WC, Fiellin DA, Merrill JO, et al. Opioid use disorder in the United States: insurance status and treatment access. Drug Alcohol Depend. 2008;94(1-3):207–213.

19. Naeger S, Mutter R, Ali MM, Mark T, Hughey L. Post-Discharge Treatment Engagement Among Patients with an Opioid-Use Disorder. J Subst Abuse Treat. 2016;69:64–71.

20. CDC. Drug Overdose Death. 2018.

21. Health MDoP. An Assessment of Fatal and Nonfatal Opioid Overdoses in Massachusetts (2011 - 2015). 2017.

22. Wu LT WG, Yang C, Blazer DG. How do prescription opioid users differ from users of heroin or other drugs in psychopathology: results from the National Epidemiologic Survey on Alcohol and Related Conditions. J Addict Med. 5(1):8.

23. Gnanavel S, Robert RS. Diagnostic and statistical manual of mental disorders, fifth edition, and the impact of events scale-revised. Chest. 2013;144(6):1974.

24. Medicine ASoA. The National Practice Guideline: For the Use of Medications in the Treatment of Addiction Involving Opioid Use. 2015.

